# Effectiveness of Paxlovid - a review

**DOI:** 10.1101/2023.01.13.23284506

**Authors:** Sydney Paltra, Tim Conrad

## Abstract

Paxlovid is an oral treatment for mild to moderate COVID-19 cases with a high risk for severe course of the disease. For this review, we have performed a comprehensive literature review. We present a summary of currently available data on Paxlovid’s ability to reduce the risk of progressing to a severe disease state. Our findings can be concluded as follows: data from the time when the Delta-variant was dominant shows that Paxlovid reduced the risk of hospitalization or death by 87.8% for unvaccinated, non-hospitalized high-risk individuals. Data from the time when the Omicron variant was dominant found decreased risk reductions, varying between 41% and 46%, combining various vaccination statuses. However, one study, which differentiated by age, found that the administration of Paxlovid reduced the risk of hospitalization by 67% for individuals aged 65 and older, but only by 27% for individuals aged 40-64. From the available data, one can conclude that Paxlovid cannot substitute vaccination, but its low manufacturing cost as well as its easy administration make it a valuable tool in fighting COVID-19, especially for countries with a low vaccination rate.

## 1 Introduction

For the past two and a half years, health officials have faced challenges of unknown dimensions due to the COVID-19 pandemic. As of September 2022, the pandemic has been responsible for more than 619 million cases and more than 6.5 million deaths world-wide [[20]]. Apart from the enormous burden on health care systems, societies have also faced psychological and socio-economical challenges [[27]; [13]; [15]; [23]], often affecting minorities the most [[14]; [26]]. Hence, it is of great interest to appropriately treat the infected and control the disease spread.

In fighting the pandemic, a lot of hope has been placed on the development of effective vaccines. Currently, six vaccines (two mRNA vaccines BNT162b2 and mRNA-1273 (and their respective Omicron updates), two vector vaccines ChAdOx1 and Ad26.COV2.S, protein-based NVX-CoV2373 and inactivated, adjuvanted VLA2001) have been authorized for use in the EU [[7]], six in the UK [[19]] and four in the USA [[3]]. A recent study [[31]] has estimated vaccinations to have saved between 14 and 20 million lives during the first year of the vaccine roll-out. But vaccine roll-out differs greatly between developed and developing countries, leading to 73% of people in the EU [[5]], but only around 27% of people in Africa [[1]] having completed a primary vaccination course. Additionally, the vaccines’ effectiveness wanes over time, and even when offered the vaccine, not everyone medically can or wants to receive the vaccine. Consequently, not all hope should be placed on vaccines and a lot of scientific effort has also been put towards the development of antiviral medication. Among the currently available treatments are:

- Remdesivir (Veklury) [[28]; [24]] was the first available treatment for COVID-19. Its administration should start as soon as possible, but must start within the first seven days of showing symptoms. Additionally, Remdesivir must be given intravenously and the length of the treatment depends on the patient’s state of disease.
- Molnupiravir [[32]], in comparison to previous treatments, allows oral intake at home, which facilitates administration. Molnupiravir was authorized in the UK in November 2021 and by the FDA in December 2021. Its administration must begin within the first five days of showing symptoms, and Molnupiravir should be taken every 12 hours over the course of five days [[17]].

Another novel treatment for mild to moderate COVID-19 cases, which allows oral intake at home, is the antiviral Paxlovid developed by Pfizer. This review aims to present the current state of literature on mainly a) Paxlovid’s efficacy in preventing progression to a more severe disease state, but also on b) whether this efficacy depends on strain, age, and vaccination status. We begin in Section 2 by describing the dosage, mode of action and potential side effects of Paxlovid before briefly introducing our methods in Section 3. In the Review section (Section 4) we present six studies (the clinical results and five subsequent publications) examining Paxlovid’s effectiveness in preventing progression to severe disease as well as its potential dependency on age, strain, and vaccination status. Finally, the Discussion section (Section 5) raises additional factors which should be taken into consideration when evaluating Paxlovid’s safety and adequacy of prescription.

## 2 Paxlovid (ritonavir and nirmatrelvir)

Paxlovid is an antiviral medicine for the treatment as well as post-exposure prophylaxis of COVID-19, consiting of nirmatrelvir tablets co-packaged with ritonavir tablets. Nirmatrelvir blocks one of SARS-CoV-2’s main polyprotein protease enzymes, M^pro^ (also referred to as 3CL protease) making it unable to bind the polyprotein. In consequence, the virus cannot replicate. Ritonavir, a protease inhibitor, is utilized as a pharmacokinetic enhancer for nirmatrelvier, curbing its breakdown in the liver and therefore allowing nirmatrelvier to reach higher systemic concentrations and dissolve slower [[8]].

Paxlovid’s recommended dosage consists of two 150mg tablets of nirmatrelvier and one 100mg tablet of ritonavir, which are to be taken together orally. The tablets are supposed to be taken over the course of five days, twice a day [[9]; [21]]. It is advised to complete the five-day treatment, but at the same time Paxlovid is not authorized for use for longer than five consecutive days. Possible side effects of Paxlovid include, but are not limited to, an impaired sense of taste, diarrhea, high blood pressure and muscle aches. Taking Paxlovid in combination with other drugs may result in drug interactions. As ritonavir may cause liver damage, Paxlovid is not recommended for patients with severe kidney or liver impairment. Additionally, patients with moderate renal impairment should be prescribed a decreased dose [[9]].

## 3 Methods

In early September 2022, we searched PubMed and MedRxiv for publications that contain at least one of the terms: “Paxlovid”, “Nirmatrelvir” and “Ritonavir” in the title and/or abstract. This resulted in 140 publications with a publication date up until August 31, 2022 which we then considered further by scanning the abstract for relevant content. Publications for which no abstract was provided as well as publications whose main text was not available in English were automatically excluded. Included were publications which discussed Paxlovid’s effectiveness in reducing the risk of progressing to a more severe disease state. Hereby, effectiveness was studied through clinical or cohort studies or in form of a retrospective analysis of data from a health care provider/health care providers. This process resulted in a total of six studies that were included in this review.

## 4 Review

The first study considered in this review is the phase 2–3, double-blind, randomized, placebo-controlled trial by [12]. Patients were recruited from (in decreasing order, based on the number of sites involved) the United States, Bulgaria, South Africa, Brazil, India, Mexico, Ukraine, Turkey, Japan, Spain, Russia, Argentina, Colombia, Poland, South Korea, Hungary, Taiwan, Malaysia, Czech Republic, Thailand and Puerto Rico while Delta was the dominant variant. Eligible patients had to be at least 18 years old with a confirmed SARS-CoV-2 infection (the authors do not specify if this solely includes PCR-confirmed individuals or both rapid antigen and PCR-confirmed individuals) and symptom onset of no more than five days before randomization, have at least one sign or symptom of COVID-19 on the day of randomization and at least one characteristic or coexisting condition associated with high risk of progression to severe COVID-19^1^. Key exclusion criteria were prior receipt of any SARS-CoV-2 vaccine or convalescent plasma, previous confirmed COVID-19 infection or hospitalization for COVID-19 and anticipated need for hospitalization within 48 hours of randomization. Additional exclusion criteria are listed in their supplementary appendix. Here, 2,246 patients underwent randomization, of which 1,120 received Paxlovid and 1,126 received a placebo every 12 hours for five days (10 doses total). The final analysis at the end of the 28 day follow-up period included only patients who had commenced treatment within five days of symptom onset. Said analysis concluded that 8 of 1039 patients (0.77%) in the treated group and 66 of 1046 (6.31%) in the placebo group were hospitalized for COVID-19 or died from any cause, which corresponds to an 87.8% relative risk reduction. Subgroup analyses were performed and can be found in the supplementary appendix.

[11] searched the TriNetX^2^ research network for data for the preprint. The authors included solely vaccinated patients over the age of 18, who developed COVID-19 at least one month after their vaccination and between December 1, 2021 and April 18, 2022. Their key exclusion criteria were treatment with monoclonal antibodies, convalescent plasma or molnupiravir. Patients who received Paxlovid more than five days after their diagnosis were also excluded as well as patients requiring initial hospitalization. They identified 111,588 non-hospitalized patients during the study period, of which 1,131 received Paxlovid. Using propensity score matching they defined the treated and the control cohort, consisting of 1,130 patients each. At a follow-up 30 days after diagnosis the studied primary outcomes were all-cause emergency room (ER) visit, hospitalization or death. The relative risk reduction for all composite outcomes was 45%, while the authors furthermore identified a relative risk reduction of 41% for ER visits, a relative risk reduction of 60% for hospitalization and finally a 100% relative risk reduction for death.

For a preprint by [4], data from a large health care system, which provides care for around 1.5 million people in Massachusetts and New Hampshire (USA), was accessed. Individuals were included in this study if a) they were over 50 years old, b) if they received a COVID-19 diagnosis between January 1, 2022 and May 15, 2022 and c) if they did not test PCR positive in the 90 days before their diagnosis. The dominant variant in Massachusetts and New Hampshire at that time was first Omicron BA.1 and then Omicron BA.2, and 30,322 non-hospitalized adults, of which 87.2% were vaccinated, were considered. Of these, patients who received other approved treatments for early COVID-19 (monoclonal antibodies, molnupiravir or remdesivir) as well as patients who are taking common medications where coadministration with nirmatrelvir plus ritonavir is not advised were excluded. Additionally, patients who were first diagnosed at the time of hospital admission or death were only included in the development of testing weights. Consequently, 30,322 outpatients who were eligible for treatment with Paxlovid were considered. Of these, 6,036 (19.9%) were prescribed Paxlovid, while 24,286 (80.1%) were not. The considered key outcome was hospitalization within 14 days of infection. Of the patients prescribed Paxlovid 40 (0.66%) were hospitalized, while among the patients not prescribed Paxlovid 232 (0.96%) were hospitalized. In consequence, the authors computed an adjusted risk ratio of 0.55, i.e. a reduction of 45% for the treated group. In addition to the main analysis, subgroup analyses were performed^3^.

Using data from Clalit Health Services database^4^, but also from the Israeli Ministry of Health COVID-19 database, a study was published by [18]. During the study period, Omicron BA.1 was the dominant variant in Israel, but during the final study weeks Omicron BA.2 gained momentum. In the study, the authors included adults aged 18 and older with a first-ever positive COVID-19 test (including PCR or antigen tests), which was performed between January 1, 2022 and February 28, 2022. Additionally, these patients had to have at least one comorbidity or condition associated with high risk for severe COVID-19^5^. Exclusion criteria were the use of medications that were contraindicated for use with Paxlovid, an estimated glomerular filtration rate < 30 mL/minute/1.73 m^2^, dialysis, weight < 40 kg, or pregnancy. Patients who were treated with molnupiravir and patients who received Paxlovid more than five days following their positive test date were also excluded, as well as patients whose first positive SARS-CoV-2 test was performed during hospitalization or on the same day of hospital admission. The follow-up ended a) 28 days after diagnosis, b) with the occurrence of severe COVID-19 or death or c) on March 10, 2022, whichever came first. The authors included 180,351 patients of which 4,737 (2.6%) received Paxlovid and of which 135,482 (75.1%) were vaccinated. The considered outcome was the composite of severe COVID-19 or mortality. Using a multivariable Cox regression model, the authors found that Paxlovid was independently associated with a significantly decreased risk for the composite of severe COVID-19 or mortality, with a hazard ratio (HR) of 0.54, and furthermore that being vaccinated was also associated with significantly decreased risk (HR 0.20).

A second study using data from the Clalit Health Services database in Israel, published by [2], considered the period from January 9, 2022 until March 10, 2022, when Omicron was the dominant variant in Israel. Here, to be eligible for consideration, patients must have tested positive by February 24, 2022. Furthermore, for patients to be considered eligible for Paxlovid prescription they had to be at least 40 years old, have a confirmed COVID-19 infection (again, the authors do not specify whether this solely includes PCR-positive individuals or also rapid antigen positive individuals), considered high-risk for severe disease^6^ and determined eligible for Paxlovid therapy. Excluded from this study were patients residing in a long-term care facility, patients who were hospitalized during the study period but before a positive SARS-CoV-2 test result and patients treated with molnupiravir. A patient’s follow-up either ended a) 35 days after diagnosis or b) with the end of the study period or c) if censored due to hospitalization/death, whichever came first. In this study, 109,213 participants were considered, and the authors report results for two separate age groups: 40-64 and 65 and older. Of the 42,819 eligible patients aged 65 years and above, 2,504 (5.8%) were treated with Paxlovid. Using a multivariate Cox proportional-hazards regression model, the authors found an adjusted hazard ratio of 0.33 for hospitalization and an adjusted HR of 0.19 for death. For people aged 40-64 they found an adjusted HR of 0.78 (hospitalization) and 1.64 (death) respectively.

The most recent preprint considered in this review was published by [33], who performed a retrospective cohort study on hospitalized patients in Hong Kong. The authors considered patients who were diagnosed between February 26, 2022 and April 26, 2022 with COVID-19, while Omicron BA.2 was the dominant variant in Hong Kong. Taking data from 40,776 hospitalized, but non-oxygen-dependent patients into account, they matched oral antiviral users and controls using propensity-score matching in an 1:1 ratio. In consequence, 1,856 molnupiravir (Mol) users, 890 Paxlovid(Pax) users and 2,746 control patients over a mean follow-up of 41.3 days were included to study mortality, composite outcome of disease progression, individual outcomes as well as the time to achieve lower viral burdens of cycle threshold. The authors concluded that antiviral users had a significantly lower risk of all-cause mortality (Mol HR=0.48, Pax HR =0.24), composite outcome of disease progression (Mol HR=0.69, Pax HR=0.57) and need for oxygen therapy (Mol HR= 0.69, Pax HR = 0.73). Additionally, they found the time to achieve a lower viral burden to be significantly shorter among antiviral users. It is to be noted that since this study only examines hospitalised patients, it is only partially comparable to the previously listed studies.

**Table 1:** Efficacy results from the six studies presented in this review. Here, an event is the occurrence of the studied outcome. [33] consider hospitalized patients, all remaining studies consider non-hospitalized patients.

| Publication | Studied Outcome | Treated Group<br>(Events) | Untreated/<br>Placebo Group<br>(Events) | Relative Risk/<br>Hazard Ratio |
| --- | --- | --- | --- | --- |
| Hammond et al. [12] | Hospitalization or Death | 1039 (8) | 1046 (66) | RR 0.122 |
| Ganatra et al. [11] | Emergency room visit | 1130 (82) | 1130 (142) | RR 0.59 |
|  | Hospitalization | 1130 (10) | 1130 (23) | RR 0.40 |
|  | Death | 1130 (0) | 1130 (10) | RR 0.00 |
|  | Composite of the above | 1130 (89) | 1130 (144) | RR 0.55 |
| Dryden-Peterson et al. [4] | Hospitalization | 6036 (40) | 24,286 (232) | RR 0.55 |
| Najjar-Debbiny et al. [18] | Hospitalization or Death | 4737 (39) | 175,614 (903) | HR 0.54 |
| Arbel et al. [2] | Hospitalization (65+) | 2504 (14) | 40,315 (762) | HR 0.33 |
|  | Death (65+) | 2504 (2) | 40,315 (151) | HR 0.19 |
|  | Hospitalization (40-64) | 1435 (9) | 64,959 (334) | HR 0.78 |
|  | Death (40-64) | 1435 (1) | 64,959 (13) | HR 1.64 |
| Wong et al. [33] | Death | 890 (32) | 890 (92) | HR 0.34 |
|  | Invasive mechanical ventilation | 890 (6) | 890 (6) | HR 0.97 |
|  | ICU admission | 890 (0) | 890 (1) | HR NA |
|  | Needing Oxygen Therapy | 890 (79) | 890 (102) | HR 0.73 |
|  | Composite of the above | 890 (101) | 890 (173) | HR 0.57 |

## 5 Discussion

In this review, we have found that a) the presented studies compute different levels of effectiveness for Paxlovid. This may be traced back to b) the different study periods (the initial clinical study was performed while Delta was the dominant variant, while subsequent studies were performed while Omicron was the dominant variant) and different study populations (the clinical study included only unvaccinated individuals, but subsequent studies included both vaccinated and unvaccinated individuals). Here, further subanalyses are necessary to single out whether age, strain or vaccination status are the cause of the difference in effectiveness. Furthermore, the variation in study design only allows for a limited comparison. Still, Paxlovid has proven to remain effective for current variants both in vivo and in vitro [[29]]. But, it shall be noted that this review solely focuses on Paxlovid’s ability to reduce the risk of progressing to a more severe disease state. When evaluating its usefulness, further factors should be taken into consideration: When practitioners or pharmacies have to judge the adequacy of prescription, they should additionally contemplate a) Paxlovid’s effect on developing Long Covid, b) potential medical interactions, and c) potential rebound effects. Paxlovid’s positive effect on the probability of developing Long Covid has been the topic of a recent publication [[34]], while others discussed potential medication interaction [[10]; [16]; [22]]. Note, that in most of the studies we have discussed, patients who are taking medications for which coadministration with nirmatrelvir plus ritonavir is not advised were excluded. For these patients, Paxlovid is not an option and one should concentrate on alternatives to protect these members of society. Furthermore, potential rebound effects [[6]]; [[30]; [25]] were not part of this review. The frequency and severity of these effects should be examined further to be able to adequately judge Paxlovid’s effectiveness. Altogether, from the available literature we come to the conclusion that Paxlovid is a valuable tool, especially in countries with a low vaccination rate, but it cannot replace vaccines and is not a panacea in the fight of the COVID-19 pandemic.

## Data Availability

No data was produced in the present work.

## Footnotes

1 These characteristics/conditions are: *≥*60 years of age; BMI *>* 25 kg/m^2^, cigarette smoking; immunosuppressive disease (including HIV infection with CD4 cell count *<* 200 mm^3^ and VL *<* 400copies/mL) or prolonged iatrogenic immunosuppression; chronic lung, cardiovascular, kidney, or sickle cell disease; hypertension; diabetes; cancer; neurodevelopmental disorders or other medically complex conditions; or medical-related technological dependence.

2 TriNetX is a clinical Data Management Software.

3 For example, they consider the age groups 50-64, 65-79 and 80+ separately and find a relative risk of 0.33, 0.60 and 0.85. Furthermore, for vaccinated patients they find a relative risk of 0.72 and for not fully vaccinated patients (they do not specify if this includes unvaccinated patients) the relative risk is 0.16. For results for the subgroups based on socioeconomic vulnerability of zip code, race and ethnicity, vaccination timing, comorbidity score, immuncompromise status and body mass index see figure 3 in the preprint’s appendix.

4 CHS provides healthcare for around 4.7 million people, half of the Israeli population.

5 Including age *≥*60 years, body mass index (BMI) *≥*30 kg/m^2^, diabetes, hypertension, cardiovascular disease, chronic liver disease, chronic lung disease, chronic kidney disease, neurological disease, immuno-suppression, and malignancy. For the full list, see https://www.cdc.gov/coronavirus/2019-ncov/hcp/clinical-care/underlyingconditions.html.

6 This is based on a score developed by the Clalit Health Services including the factors age, vaccination status, immunsuppression, hospitalization events in the previous 3 years, body mass index, COPD, heart disease, vascular disease, cerbrovascular, renal, hepatic or neurological disease, active malignancy, diabetes, organ or bone marrow transplant, previous splenectoms, AIDS patient or HIV carrier and treatment with immunosuppressants or steroid treatment at least twice in the previous year.

