## Supplemental Table 1, 2, and 3 for "Effectiveness of Paxlovid - a review"

### Supplementary Material

The following section contains three tables: The first table contains the preprints and papers, which met the inclusion criteria and were considered in our review. The second table contains the preprints and papers, which were included in our study, but not the review itself and the final table includes all papers and preprints, whose title and abstracts were scanned, but which did not meet the inclusion criteria.

#### Used

| **Author(s)** | **Title** | **Type of publication & journal** |
|---|---|---|
| (Arbel *et al.*, 2022) | Oral Nirmatrelvir and Severe Covid-19 Outcomes During the Omicron Surge | Preprint |
| (Dryden-Peterson *et al.*, 2022) | Nirmatrelvir plus ritonavir for early COVID-19 and hospitalization in a large US health system | Preprint |
| (Ganatra *et al.*, 2022) | Oral Nirmatrelvir and Ritonavir in Non-hospitalized Vaccinated Patients with Covid-19 | Published in *Clinical Infectious Diseases* |
| (Hammond *et al.*, 2022) | Oral Nirmatrelvir for High-Risk, Nonhospitalized Adults with Covid-19 | Published in *New England Journal of Medicine* |
| (Najjar-Debbiny *et al.*, 2022) | Effectiveness of Paxlovid in Reducing Severe Coronavirus Disease 2019 and Mortality in High-Risk Patients | Published in *Clinical Infectious Diseases* |

| | | |
|---|---|---|
| (Wong *et al.*, 2022) | Real-world effectiveness of early molnupiravir and nirmatrelvir/ritonavir among hospitalized, non-oxygen-dependent COVID-19 patients on admission during Hong Kong's Omicron BA.2 wave: an observational study | Preprint |

**Table S1:** Publications which were considered in our review.

### Used, but not in review

| Author(s) | Summary | Used in section |
|---|---|---|
| (Epling *et al.*, 2022) | Examination (clinically, virologically and immunologically) of seven patients who experienced rebound symptoms. Six experienced rebound symptoms after taking Paxlovid, the final patient did not receive the treatment. | Discussion |
| (Extance, 2022) | Short sections on who is eligible for treatment with Paxlovid, how does Paxlovid work, what peer reviewed evidence is there for Paxlovid, which countries are using Paxlovid, how much does Paxlovid cost, what is the potential for this drug. | Paxlovid (Ritonavir and Nirmatrelvir) |
| (Fishbane, Hirsch and Nair, 2022) | Argue that Paxlovid might harm transplant patients as there is a chance of medication interaction and that the ritonavir component of paxlovid might pose a safety risk for these patients | Discussion |
| (L. Wang, Volkow, *et al.*, 2022) | Discussion of if and how rebound symptoms after taking Paxlovid differ between Omicron BA.5 and | Discussion |

| Author(s) | Summary | Used in section |
|---|---|---|
| | Omicron BA.2.12.1 infected patients | |
| (Marzolini *et al.*, 2022) | Review which summarized the effects of ritonavir on drug disposition and debates what influences the probability of drug-drug interaction when taking Paxlovid. | Discussion |
| (Prikis and Cameron, 2022) | Discusses side-effects of Paxlovid in a single kidney transplant patient, whos treatment with Paxlovid had to be interrupted and who suffered acute kidney injury from taking the drug | Discussion |
| (Rubin, 2022) | No abstract available on Pubmed, published under “Medical News & Perspectives”, discussing rebound effects potentially associated with Paxlovid treatment. | Discussion |
| (Vangeel *et al.*, 2022) | Assessment of the in vitro antiviral effect of GS-441524, remdesivir, EIDD-1931, molnupiravir and nirmatrelvir against the various SARS- CoV-2 VOCs, including Omicron. | Discussion |
| (Wen *et al.*, 2022) | Review of efficacy and safety of the three antiviral treatments molnupiravir, fluvoxamine and Paxlovid. Published, when only Paxlovid’s clinical results were available. | Introduction |

**Table S2:** Publications which were considered in our paper, but not in the review itself.

### Unused Table

| Author(s) | Summary | Reason for Exclusion |
|---|---|---|
| (Abdelrahim *et al.*, 2022) | Discussing Thymoquinone and other natural products as possible treatments for COVID-19. | Does not meet the inclusion criteria. |
| (Alshanqeeti and Bhargava, 2022) | Case discussion of two patients who were prescribed Paxlovid and experiences a rebound of COVID-19 | Does not meet the inclusion criteria. |
| (Alvarado *et al.*, 2022) | Paxlovid binds non-covalently at regions other than the catalytic sites with energies stronger than reported, namely to $mM_{pro}$. | Does not meet the inclusion criteria. |
| (Ashour *et al.*, 2022) | Review which presents the efficacy of repurposed drugs for COVID-19. Discussion of clinical trials, combination therapies and novel methods followed for treatment. Written when Paxlovid was undergoing Phase III studies. | Does not meet the inclusion criteria. |
| (Atluri, Aimlin and Arora, 2022) | Debating evidence from pivotal trials that led to the approval of effective COVID-19 therapeutics and categorizing them as effective outpatient and inpatient management strategies. | Does not meet the inclusion criteria. |
| (Azanza *et al.*, 2022) | Description of drugs that contraindicated and/or should (not) be used carefully when also taking Paxlovid based on fact sheets by the Spanish Agency for Medicines and Health Products. | Does not meet the inclusion criteria. |
| (Bartha *et al.*, 2022) | Proposition of a hybrid multiscale mathematical approach to assess Paxlovid. | Does not meet the inclusion criteria. |
| (Ben Hlima *et al.*, 2022) | Usage of techniques of structure modeling, in silico | Does not meet the inclusion criteria. |

| Author(s) | Summary | Reason for Exclusion |
|---|---|---|
| | docking and pharmacokinetics prediction to test compounds from algae for their ability to inhibit SARS-CoV-2's protease Mpro. | |
| (Berar Yanay *et al.*, 2022) | No abstract available on pubmed. | |
| (Birabaharan and Martin, 2022) | Discussion of the case of a male veteran reporting rebound symptoms after Paxlovid treatment. The patient was found to be hypoxic with pulmonary emboli. | Does not meet the inclusion criteria. |
| (Borio, Bright and Emanuel, 2022) | No abstract available on pubmed. | Abstract unavailable. |
| (Brooks, Song and Sultan, 2022) | No abstract available on pubmed. | Abstract unavailable. |
| (Burki, 2022) | No abstract available on pubmed. | Abstract unavailable. |
| (Buxeraud, Faure and Fougere, 2022) | The English abstract solely states that Paxlovid is an available treatment for COVID-19 infections, administered orally and to people at high risk for severe disease. | Only the abstract was available in English. |
| (Callaway, 2022) | No abstract available on pubmed. | Abstract unavailable. |
| (Catlin *et al.*, 2022) | Written when Paxlovid was still under development, embryo-fetal development studies in rats and rabbits to assess clinically relevant risks when prescribing Paxlovid to males and females in reproductive age. | Does not meet the inclusion criteria. |
| (Cerón-Carrasco, 2022) | Aim to critically assess benefits and shortcomings of using molecular models for drug repurposing when trying to develop effective | Does not meet the inclusion criteria. |

| Author(s) | Summary | Reason for Exclusion |
|---|---|---|
| | COVID-19 treatments.. | |
| (Chen, Chang and Wei, 2022) | No abstract available on pubmed. | Abstract unavailable. |
| (Choi *et al.*, 2022) | Discussing panax ginseng's (a medical plant) ability to control cytokine storm in COVID-19 | Does not meet the inclusion criteria. |
| (Collaborative *et al.*, 2022) | Identification of patients potentially eligible for treatment with Paxlovid in the UK. In consequence, assessment of the coverage of new treatments among these patients with the conclusion that there were variants in coverage between key clinical, geographic and demographic groups. | Does not meet the inclusion criteria. |
| (Coulson *et al.*, 2022) | No abstract available on pubmed. | Abstract unavailable. |
| (Couzin-Frankel, 2021) | No abstract available on pubmed. | Abstract unavailable. |
| (Dai *et al.*, 2022) | For 36 mRNA-vaccinated and Omicron-infected individuals viral kinetics were measured. 11 of the 36 were treated with Paxlovid and treatment was associated with larger incidence of viral rebound. | Does not meet the inclusion criteria. |
| (de Oliveira *et al.*, 2022) | Report of simulation of H172Y mutation on Mpro's structure leading to decreased structural stability and binding affinity. | Does not meet the inclusion criteria. |
| (Deo *et al.*, 2022) | Evaluation of the incidence of viral and symptom rebound in untreated mild to moderate COVID-19 outpatients. | Does not meet the inclusion criteria. |
| (Drożdżal *et al.*, 2021) | Review presenting the progress in clinical trials concerning the effectiveness | Does not meet the inclusion criteria. |

| Author(s) | Summary | Reason for Exclusion |
|---|---|---|
| | of treatments for COVID-19. Written when solely the results from Paxlovid's clinical trials were available. | |
| (Rubin, Baden and Morrissey, 2022) | No abstract available on pubmed. | Abstract unavailable. |
| (Eng *et al.*, 2022) | Presenting the preclinical disposition, metabolism and potential drug-drug interaction of nirmatrelvir. | Does not meet the inclusion criteria. |
| (Feingold, 2022) | Discussion of levels of total cholesterol, LDL-C, HDL-C, and apolipoprotein B and A-I levels in patients with COVID-19 infections. | Does not meet the inclusion criteria. |
| (Feng *et al.*, 2022) | Discussing the potential of Yindan Jiedu granules as a treatment for COVID-19. Comparison of Yindan Jiedu granules and Paxlovid when treating COVID-19 patients. | Does not meet the inclusion criteria. |
| (Fernando *et al.*, 2022) | Present how Pfizer has successfully improved its research and development (R&D) productivity between 2010 and 2020. | Does not meet the inclusion criteria. |
| (Ferrara *et al.*, 2022) | Presentation of literature on monotherapy use of Paxlovid and monotherapy use of remdesivir and discussion of the hypothesis of using nirmatrelvir and remdesivir to increase efficacy of COVID-19 treatment. | Does not meet the inclusion criteria. |
| (Gandhi, Malani and Del Rio, 2022) | No abstract available on pubmed. | Abstract unavailable. |
| (García-Lledó *et al.*, 2022) | Provides a review of currently available COVID-19 treatments. As this was published in December 2021, it only include the clinical trials for Paxlovid. | Does not meet the inclusion criteria. |

| Author(s) | Summary | Reason for Exclusion |
|---|---|---|
| (Gold *et al.*, 2022) | Data from December 2021 until May 2022 were analyzed to describe oral antiviral treatment prescription dispensing overall and by week, stratified by zip code and social vulnerability. | Does not meet the inclusion criteria. |
| (Greasley *et al.*, 2022) | Evaluation of in vitro potency of normatrelvir against the Mpro of currently circulating (and previous) variants of concern. Their in vitro data suggests that Paxlovid has the ability to inhibit SARS-CoV-2 replication, even vor VOCs like Omicron. | Does not meet the inclusion criteria. |
| (Halford, 2022) | No abstract available on pubmed. | Abstract unavailable. |
| (Ho *et al.*, 2022) | Retrospective analysis of benefits and limitations of (previous) treatments for COVID-19. Calls Paxlovid a promising treatment, but only presents clinical results. | Does not meet the inclusion criteria. |
| (Hong *et al.*, 2022) | People with cystic fibrosis (CF) are at risk for drug-drug interaction when being treated with Paxlovid. Simulation of coadministration of elexacaftor-tezacaftor-ivacaftor and Paxlovid to examine these potential interactions. | Does not meet the inclusion criteria. |
| (Huang *et al.*, 2022) | Case report of a severe aplastic anemia child who was successfully treated with Paxlovid. | Does not meet the inclusion criteria. |
| (Hung *et al.*, 2022) | Written before Paxlovid was authorized and before clinical results were available. Hence, discussing the promising antiviral effect of nirmateelvir, while also | Does not meet the inclusion criteria. |

| Author(s) | Summary | Reason for Exclusion |
|---|---|---|
| | naming (then) unresolved concerns. | |
| (Islam *et al.*, 2022) | Evaluation of drugs authorized in the US for treating COVID-19. For Paxlovid's effectiveness, solely the clinical results are discussed. | Does not meet the inclusion criteria. |
| (Javaux and Ader, 2022) | Discussing medical management (but excluding intensive care management) of COVID-19. | Article only available in French. |
| (Joyce, Hu and Wang, 2022) | Review to present the potential of nirmatrelvir when treating COVID-19, discussing its history of rational design, its target selectivity, synthesis and drug resistance. | Does not meet the inclusion criteria. |
| (Katella, 2022) | News article introducing the most important Paxlovid-related information for the general public and patients. | Does not meet the inclusion criteria. |
| (Kozlov, 2022) | No abstract available on pubmed. | Abstract unavailable. |
| (Kuehn, 2022) | No abstract available on pubmed. | Abstract unavailable. |
| (L. Wang, Berger, *et al.*, 2022) | Examination of rates and relative risks of COVID-19 rebound in patients treated with Molnupiravir or Paxlovid during January-June 2022 | Does not meet the inclusion criteria. |
| (Lamb, 2022) | Summary of the milestones during the development of normatrelvir plus ritonavir leading to its first authorizations and approval for the treatment of COVID-19 | |
| (Ledford and Maxmen, 2022) | No abstract available on pubmed. | Abstract unavailable. |

| **Author(s)** | **Summary** | **Reason for Exclusion** |
|---|---|---|
| (Ledford, 2022) | No abstract available on pubmed. | Abstract unavailable. |
| (Lee *et al.*, 2022) | Discussion of potential nirmatrelvir escape mutations from emerging variants of SARS-CoV-2 and exploration of the mutational landscape of $M_{pro}$. | Does not meet the inclusion criteria. |
| (Lemaitre, Budde, *et al.*, 2022) | Discussion of drug interactions of Paxlovid and immunosuppressant drugs and provision of general recommendations for therapeutic drug monitoring when co-administering the two. | Does not meet the inclusion criteria. |
| (Lemaitre, Grégoire, *et al.*, 2022) | Providing recommendations on behalf of the national French society of pharmacology for possible drug-drug interactions between Paxlovid and other commonly used drugs. | Does not meet the inclusion criteria. |
| (Li *et al.*, 2022) | No abstract available on pubmed. | Abstract unavailable. |
| (Lieber and Plemper, 2022) | Introduction of a potential future treatment, the orally available ribonucleoside analog 4'-fluorouridine (4'-FIU). Furthermore, reviewing currently approved and emerging medicines against COVID-19. | Does not meet the inclusion criteria. |
| (Chengyu Liu *et al.*, 2022) | Discussing statin use in the context of COVID-19, coming to no evidence suggesting interference between statins and COVID-19 vaccines. But, simultaneous stantis and Paxlovid administration may increase statin exposure and the risk of adverse effects. | Does not meet the inclusion criteria. |

| Author(s) | Summary | Reason for Exclusion |
|---|---|---|
| (Chenxi Liu *et al.*, 2022) | Development of an efficient LC-MS/MS method for simultaneously determining nirmatrelvir and ritonavir in human plasma. | Does not meet the inclusion criteria. |
| (Logue *et al.*, 2022) | Show that Apilimod and other PIKfyve inhibitors worsen disease in a COVID-19 murine model when given prophylactically or therapeutically. Abstract solely mentions Paxlovid as an available treatment. | Does not meet the inclusion criteria. |
| (Lu *et al.*, 2022) | Multicentre cohort study describing clinical characteristics, and assessing risk and protective factors for geriatric Omicron severe infections. | |
| (Mahaboob Ali *et al.*, 2022) | Discussing herbal extracts as potential treatments for COVID-19. Solely mentions that molnupiravir and Paxlovid are not widely available. | Does not meet the inclusion criteria. |
| (Mahase, 2021) | News article in BMJ, published shortly after the analysis of the phase II-III data was published. Hence, discussing and reporting the results of this analysis. | Does not meet the inclusion criteria. |
| (Malden *et al.*, 2022) | Consideration of electronic health record from a large integrated health care system in California to analyze and quantify hospital admissions and emergency department encounters related to COVID-19 during the 5-15 days after receiving Paxlovid treatment | Does not meet the inclusion criteria. |
| (Manus, 2022) | No abstract available, article in French. | |

| Author(s) | Summary | Reason for Exclusion |
|---|---|---|
| (Marzi *et al.*, 2022) | Report the discovery and description of nirmatrelvir. Comparison of effectiveness of molnupiravir and normatrelvir). Written while Paxlovid was under study in phase III of the clinical trial. | Does not meet the inclusion criteria. |
| (McDonald and Lee, 2022b) | Praxis-relevant recommendations for the usage and prescription of Paxlobid. | Does not meet the inclusion criteria. |
| (McDonald and Lee, 2022a) | No abstract available on pubmed. | |
| (McMillan, Morris and Idris, 2022) | News article published in EMBO Molecular Medicine, discussing (Chang *et al.*, 2022) | Does not meet the inclusion criteria. |
| (Mikus *et al.*, 2022) | Compilation of a list of drugs and their potentially relevant interactions when being administered simultaneously with Paxlovid as well as a list of commonly prescribed drugs for which no such interactions exist. | Does not meet the inclusion criteria. |
| (Moghadasi *et al.*, 2022) | Discussion of how ongoing virus evolution has the potential to yield variants with resistance to clinical protease inhibitors (like Paxlovid). | Does not meet the inclusion criteria. |
| (Mohapatra *et al.*, 2022) | Published when solely Paxlovid's clinical results were available. Hence, only these are discussed. | Does not meet the inclusion criteria. |
| (Mótyán *et al.*, 2022) | Review of normatrelvir's binding to SARS-CoV-2's Mpro and its potential inefficacy when novel mutations arise. | Does not meet the inclusion criteria. |
| (Ng *et al.*, 2022) | Presentation of current antiviral drugs for COVID-19 as well as potential future directions. | Does not meet the inclusion criteria. |

| Author(s) | Summary | Reason for Exclusion |
|---|---|---|
| (Nocentini, Capasso and Supuran, 2022) | Review of literature on the drug design landscape of SARS-CoV-2 $M_{pro}$ inhibitors. Mentions nirmatrelvir as one such inhibitor. | Does not meet the inclusion criteria. |
| (Ou *et al.*, 2022) | Development of a non-pathogenic system in which yeast growth is a proxy for $M_{pro}$ activity. Consequently, mutants which exhibit drug sensitivity and altered enzymatic activity can quickly be identified. | Does not meet the inclusion criteria |
| (Ouyang *et al.*, 2022) | Discussion of review of risk factors for severe covid and Pre-exposure prophylaxis (PrEP) strategies against SARS-CoV-2. Additionally, presentation of potential SARS-CoV-2 PrEP drugs | Does not meet the inclusion criteria. |
| (Parums, 2022) | Published in late 2021, reporting the current status of oral treatments for COVID-19 | Does not meet the inclusion criteria. |
| (Pavan *et al.*, 2022a) | Analysis of newly reported bat coronaviruses with regard to the similarities and differences between their 3CL protease and SARS-CoV-2. | Does not meet the inclusion criteria |
| (Pavan *et al.*, 2022b) | Analysis of the structural features of the Spike protein and the $M_{pro}$ of the SARS-CoV-2 variant XE and the closely related variants XD and XF. | Does not meet the inclusion criteria. |
| (Pawankar *et al.*, 2022) | Comparison and changes in the epidemiology, clinical profile, therapeutics and public health measures for the COVID-19 pandemic in the Asia Pacific region. Abstract mentions recent introduction of Paxlovid. | Does not meet the inclusion criteria. |

| Author(s) | Summary | Reason for Exclusion |
|---|---|---|
| (Peluso *et al.*, 2022) | Report of 4 cases from a post-COVID cohort study who received nirmatrelvir as part of clinical care and who experiences different outcomes. | Does not meet the inclusion criteria. |
| (Persad, Peek and Shah, 2022) | Published in late 2021, when the federal government rationed Paxlovid doses. Hence, this paper identifies relevant ethical principles and priority groups for access to Paxlovid. | Does not meet the inclusion criteria. |
| (Pesko *et al.*, 2022) | No abstract available on pubmed. | |
| (Phizackerley, 2022) | No abstract available on pubmed. | |
| (Priya, Basit and Bandyopadhyay, 2022) | Examine modifications of a promising peptide-based inhibitor of the spike protein, LCB3, against common mutations in the target protein. In consequence, LCB3 retains its efficacy against the spike protein. | Does not meet the inclusion criteria. |
| (Reina and Iglesias, 2022) | Discussing Paxlovid's efficacy after publication of clinical results. Article is in Spanish. | Does not meet the inclusion criteria. |
| (Ridgway *et al.*, 2022) | Discussion of a new class of sartan-like arterial antihypertensive drugs (referred to as “bisartans”) as treatment for COVID-19. Abstract notes that bisartans do not inhibit SARS-CoV-2 infection in bioassays as effectively as Paxlovid. | Does not meet the inclusion criteria. |
| (Roberts, Duncan and Cairns, 2022) | No abstract available on pubmed. | |
| (S K *et al.*, 2022) | Discussion of risk of drug interactions between treatments for COVID-19 | Does not meet the inclusion criteria. |

| Author(s) | Summary | Reason for Exclusion |
|---|---|---|
| | and drugs used in treating comorbid conditions like diabetes or cardiovascular illness. | |
| (Wang, Gelfand and Calabrese, 2022) | Summarizes diagnostic and therapeutic management of COVID-19 for outpatients. Abstract names Paxlovid was the preferred treatment for mild cases with high risk of disease progression. | Does not meet the inclusion criteria. |
| (Sakamuru, Huang and Xia, 2022) | Assessing currently available COVID-19 drugs for their potential toxicological effects and mechanisms. | Does not meet the inclusion criteria. |
| (Salerno *et al.*, 2022) | Describes clinical experience with 25 organ transplant recipients who were prescribed Paxlovid. Their results suggest that clinically significant interaction between Paxlovid and immunosuppressive agents can be reasonably managed with a standardized dosing protocol | Does not meet the inclusion criteria. |
| (Sánchez Fabra and Herrero Jordán, 2022) | No abstract available on pubmed. | |
| (Saravolatz, Depcinski and Sharma, 2022) | Review of molnupiravir and Paxlovid with regard to their mechanisms of action, their antiviral activity, pharmacokinetics, drug interactions and clinical experience including trials, adverse events, recommended indications and formulary considerations. | Does not meet the inclusion criteria |
| (Sathish *et al.*, 2022) | Providing vivo safety assessments of Paxlovid by studies in rats and monkeys. | Does not meet the inclusion criteria. |
| (Schöning *et al.*, 2022) | Simulation of molnupiravir | Does not meet the inclusion |

| Author(s) | Summary | Reason for Exclusion |
|---|---|---|
| | treatment to judge effectiveness of antiviral therapy in highly-transmissible variants. | criteria. |
| (Schwartz, 2022) | No abstract available on pubmed. | |
| (Secretan *et al.*, 2022) | Discussing the intrinsic stability of Nirmatrelvir and the degrading products formed under forced conditions. | Does not meet the inclusion criteria. |
| (Service, 2022) | No abstract available on pubmed. | |
| (Setz *et al.*, 2022) | Introduce the ability of the combination of pamapimod and pioglitazone to inhibit SARS-CoV-2 replication in vitro. Hence, this combination is a potential treatment of COVID-19 and is evaluated (at the time of writing) in a phase II clinical study. | Does not meet the inclusion criteria. |
| (Singh *et al.*, 2022) | Provides practical clinical guidelines for using molnupiravor in COVID-19 patients. | Does not meet the inclusion criteria. |
| (Stifani *et al.*, 2022) | Discussing contraceptive care in times of the Pandemic. Abstract mentions that combined hormonal contraceptive users who take Paxlovid should consider an additional contraceptive method for the duration of Paxlovid treatment. | Does not meet the inclusion criteria. |
| (Sun *et al.*, 2022) | No abstract available on pubmed. | |
| (Tang *et al.*, 2022) | Development and validation of a method to quantify almonertinib in rat plasma to study the effects of Paxlovid on the pharmacokinetics of | Does not meet the inclusion criteria. |

| Author(s) | Summary | Reason for Exclusion |
|---|---|---|
| | almonertinib in rats. | |
| (Tanne, 2022) | No abstract available on pubmed. | |
| (Tarnawski and Ahluwalia, 2022) | Presenting the role of endothelial and vascular components as major targets for COVID-19-induced tissue injury, spreading to various organs and injury healing as well as current COVID-19 treatments. | Does not meet the inclusion criteria. |
| (Tene *et al.*, 2022) | Israelian study to determine the number of patients who could be included in a prospective Real World Evidence Study to study Paxlovid's effect on patients' outcomes as well as assessment of comparability between patients who received the treatment and patients who did not. | |
| (Tolomeo, Cavalli and Cascio, 2022) | Discussing the important role of the signal transducer and activator of transcription (STAT) 1 protein in antiviral immune response and how viruses like Ebola and SARS-CoV-2 have developed the ability to inhibit this transcription factor. | Does not meet the inclusion criteria. |
| (Traynor, 2022) | No abstract available on pubmed. | |
| (Uchikoba, Yamada and Tsuzuki, 2022) | No abstract available on pubmed. | |
| (Usher, 2022) | No abstract available on pubmed. | |
| (Viedma Martínez, Gallo Pineda and Jiménez Gallo, 2022) | Review of the indications of immunosuppresants and immunomodulators and guidance on which mild to moderate COVID-19 | Does not meet the inclusion criteria. |

| Author(s) | Summary | Reason for Exclusion |
|---|---|---|
| | patients might benefit from their use in dermatology. | |
| (Vitiello, Ferrara, *et al.*, 2022) | Review which provides an overview regarding the molecular profile of the Omicron variant, as well as its transmissibility and the remaining vaccine effectiveness. Mentioned molnupiravir and Paxlovid as oral treatments of COVID-19. | Does not meet the inclusion criteria. |
| (Vitiello, La Porta, *et al.*, 2022) | Written when the Omicron variant hat just emerged and molnupiravir and Paxlovid had only undergone clinical study. Briefly reviewing the impact of these two new oral antivirals. | Does not meet the inclusion criteria. |
| (Vuorio, Kovanen and Raal, 2022) | Discussion of drug interactions when simultaneously taking Paxlovid and cholesterol-lowering drugs. | Does not meet the inclusion criteria. |
| (Wu *et al.*, 2022) | Summary of the current progress in the structural biology of SARS-CoV-2 as well as presentation of structure-based design of Paxlovid, molnupiravor and VV116 to emphasize the importance of structure in drug development for COVID-19 | Does not meet the inclusion criteria. |
| (Y. Wang *et al.*, 2022) | No abstract available on pubmed. | |
| (Yan *et al.*, 2022) | Recruitment of 5 pediatric cases with underlying disease who were treated with Paxlovid as well as 30 age-matched patients with underlying disease who were not treated with Paxlovid as controls. Assessment of efficacy and safety of Paxlovid as well as | Does not meet the inclusion criteria. |

| Author(s) | Summary | Reason for Exclusion |
|---|---|---|
| | inter-group comparisons. | |
| (Yang *et al.*, 2022) | From evolutionary and structural standpoints possible mutations in Mpro leading to evasion of nirmatrelvir are discussed. | Does not meet the inclusion criteria. |
| (Young, Papiro and Greenberg, 2022) | Case report of a 14 year old female kidney transplant, whose COVID-19 infection was treated with Paxlovid. | Does not meet the inclusion criteria. |
| (Wang and Chan, 2022) | Cystic fibrosis patients are at increased risk for drug-drug interaction when taking Paxlovid. Here, these interactions are explored using a physiologically-based pharmacokinetic modeling approach. | Does not meet the inclusion criteria. |
| (Wang and Yang, 2022) | Letter to the editor discussing the potential of Paxlovid to treat COVID-19 | Does not meet the inclusion criteria. |
| (Zhang, 2022) | Review presenting an update on fluorinated COVID-19 drugs. Here, current knowledge of these drugs' molecular design, metabolism and pharmacokinetics as well as mechanism of action, is provided. | Does not meet the inclusion criteria. |
| (Zhu and Ang, 2022) | Review which aims to provide a compact and updated summary of pediatric COVID-19 diagnosis and management. Abstract mentions that in patients at increased risk of progression, Paxlovid (among other treatments) should be considered. | Does not meet the inclusion criteria. |

**Table S3:** Publications, which were taken into consideration, but which did not meet our inclusion criteria and were hence not included in the review.
